# Incorporating computer vision on smart phone photographs into screening for inflammatory arthritis: results from an Indian patient cohort

**DOI:** 10.1101/2024.08.19.24312283

**Authors:** Sanat Phatak, Ruchil Saptarshi, Vanshaj Sharma, Rohan Shah, Abhishek Zanwar, Pratiksha Hegde, Somashree Chakraborty, Pranay Goel

## Abstract

**Background:** Convolutional neural networks (CNNs) have been used to classify medical images; few studies use smartphone photographs that are scalable at point of care. We previously showed proof of principle that CNNs could detect inflammatory arthritis in three hand joints. We now studied a screening CNN to differentiate from controls.

**Methods:** We studied consecutive patients with early inflammatory arthritis and healthy controls, all examined by a rheumatologist (15% by two). Standardized photographs of the hands were taken using a studio box, anonymized, and cropped around joints. We fine-tuned pre-trained CNN models on our dataset (80% training; 20% test set). We used an Inception-ResNet-v2 backbone CNN modified for two class outputs (Patient vs Control) on uncropped photos. Inception-ResNet-v2 CNNs were trained on cropped photos of Middle finger Proximal Interphalangeal (MFPIP), Index finger PIP (IFPIP) and wrist. We report representative values of accuracy, sensitivity, specificity.

**Results:** We studied 800 hands from 200 controls (mean age 37.8 years) and 200 patients (mean age 49.6 years; 134 with rheumatoid arthritis amongst other diagnoses). Two rheumatologists had a concordance of 0.89 in 404 joints. The wrist was commonly involved (173/400) followed by the MFPIP (134) and IFPIP (128). The screening CNN achieved excellent accuracy (98%), sensitivity (98%) and specificity (98%) in predicting a patient compared to controls. Joint-specific CNN accuracy, sensitivity and specificity were highest for the wrist (80%, 88%, 72%) followed by the IFPIP (79%, 89%,73%) and MFPIP (76%, 91%, 70%).

**Conclusion:** Computer vision without feature engineering can distinguish between patients and controls based on smartphone photographs with good accuracy, showing promise as a screening tool prior to joint-specific CNNs. Future research includes validating findings in diverse populations, refining models to improve specificity in joints and integrating this technology into clinical workflows.

## Introduction

Artificial intelligence (AI) promises to improve the healthcare landscape, especially in automating diagnostic and screening methods. Computer vision refers to artificial neural networks used in the extraction of meaning from images with pixels and videos. (1) This is achieved via the mathematical operation called convolution. Such a convolutional neural network (CNN) can be used on various medical imaging techniques in order to recognise features, classify and segment them. (2)

CNNs’ application in medicine has progressed rapidly, since they are able to automate a variety of repetitive clinical tasks. Radiology and pathology images are most commonly used; CNNs have demonstrated the ability to classify these for a quick diagnosis. (3,4) As training datasets become larger and more nuanced, these methodologies have started to become sufficiently robust to gain widespread acceptance in clinical care. The Food and Drug Administration(FDA) approved a CNN for diabetic retinopathy grading on fundus photographs. (5) These technologies lend themselves to various conditions that can potentially be diagnosed visually.

Applications of computer vision in rheumatology are relatively few, and most have focused on radiographic or magnetic resonance (MR) images. (6) CNNs have been able to quantify and predict erosions on X-rays, thus automating the painstaking scores such as the van Der Heijde score. (7) A deep CNN could segment sacroiliac joint computed tomography (CT) images and grade sacroiliitis in axial spondyloarthritis. (8) A CNN was able to pick out rheumatoid arthritis on thermal imaging based photographs with good accuracy. (9) All these described techniques still require a patient undergoing an investigation, using specialized equipment, that limits their use to secondary or tertiary care centers or research settings. Inflammatory arthritis presents with joint pain and swelling, with visible signs of inflammation detected on inspection. We hypothesized that these features would be amenable to detection by CNNs on smartphone photographs. Our pilot data showed that CNNs could detect synovitis with reasonable accuracy in cropped images of three joints: the wrist, 2nd/ Index finger proximal interphalangeal (IFPIP) and 3rd/ middle finger PIP (MFPIP), selected based on the prevalence of involvement in the dataset. (10) Smartphones are widely available and thus this could improve point-of-care screening, triaging people with painful hand syndromes to rheumatologists as well as following up patients diagnosed with inflammatory arthritides.

We expand on previous joint-specific results by attempting a classifier CNN on uncropped hand photos to distinguish between patients with inflammatory arthritis and controls. Further the three joint-specific CNNs were trained and tested on a larger dataset incorporating controls.

## Patients and Methods

### Patients

Patients were enrolled from the rheumatology department’s outpatient clinic at the tertiary care King Edward Memorial (KEM) Hospital in Pune, India as well as from a private rheumatology practice in Pune, India. Consecutive patients with inflammatory arthritis in the hand joints, having symptoms for less than 2 years were included. Permissible clinical diagnoses included rheumatoid arthritis (RA),connective tissue diseases [systemic lupus erythematosus (SLE); Sjogren syndrome (SS); overlap, mixed and undifferentiated connective tissue diseases], chronic viral arthritis (VA), Spondyloarthritis (SA), psoriatic arthritis (PsA), Juvenile idiopathic arthritis, polymyalgia rheumatica (PMR). RA was classified according to the American College of Rheumatology/European Alliance of Associations of Rheumatology (ACR/EULAR) 2011 criteri (11), PsA according to the Classification for Psoriatic Arthritis (CASPAR) criteria (12), and SLE according to the Systemic Lupus International Collaborating Clinics (SLICC) classification criteria (13) ; all others were clinical diagnoses made by the rheumatologist. Long-standing arthritis was excluded to prevent confounding with joint deformities, as were patients with systemic sclerosis.

We recorded the age and sex of the patients, along with clinical details like duration and diagnosis of the disease; extra articular features, especially the involvement of coexistent skin lesions on the hands. The patient’s and doctor’s global perception at that clinical visit (on a visual analog scale) and results of markers of inflammation (ESR, CRP, or both) were recorded.

A trained rheumatologist (SP) conducted an independent clinical examination. Inspection, palpation, and range of motion testing in each of the 15 joints in both hands (four distal interphalangeal, four proximal interphalangeal, one interphalangeal of thumb, five metacarpophalangeal, and one wrist) were done and the presence of synovitis was recorded as binary (yes/no) opinion. Thirty patients (15%) underwent examination by two rheumatologists (SP and AZ). When there was disagreement between two rheumatologists, a consensus was reached and that was entered in the final dataset.

### Image capture and standardization

A member of trained research staff photographed both hands from the dorsal aspect in standardized conditions using a modified studio box. The dimensions of this box were 26×26×26 cm, with an inbuilt LED light system and standard background. To ensure appropriate placement of the hand, the modified studio box was kept on a desk, and patients were sat in a chair. All photographs were taken using an iPhone 11 (Apple Inc., California, United States).

### Image cropping

MediaPipe, an open-source library, located the joints of interest in the images (14). IFPIP, MFPIP and wrist joint images were cropped from the x and y coordinates extracted. Images were screened for correctness of cropping (by RS, VS) and automatic crops that did not include the entire joint (n=29) were done by hand.

Image de identification: We used random study identifiers to label photographs created by a unique identity generator. (15) Two thousand unique IDs for patients and two thousand for controls have been generated. Personally identifiable information was linked to a personal ID (IDP) while a study ID (IDS) is linked to images and clinically relevant information. The anonymisation is reversible and the key is stored securely with SP. Only anonymised images and non-identifying clinical information was shared.

### Neural Networks

We utilized pre-trained CNN models, fine-tuning them with our dataset to enhance their specificity to arthritis-related features. Transfer learning was used. The dataset was divided into training and test sets in an 80:20 ratio.

CNN 1: We used whole hand (uncropped) images of the dorsal surface of the hand. We first used an Inception-ResNet-v2 backbone CNN modified for two class outputs (Patient vs control) (Fig 1A)

**Figure 1:**
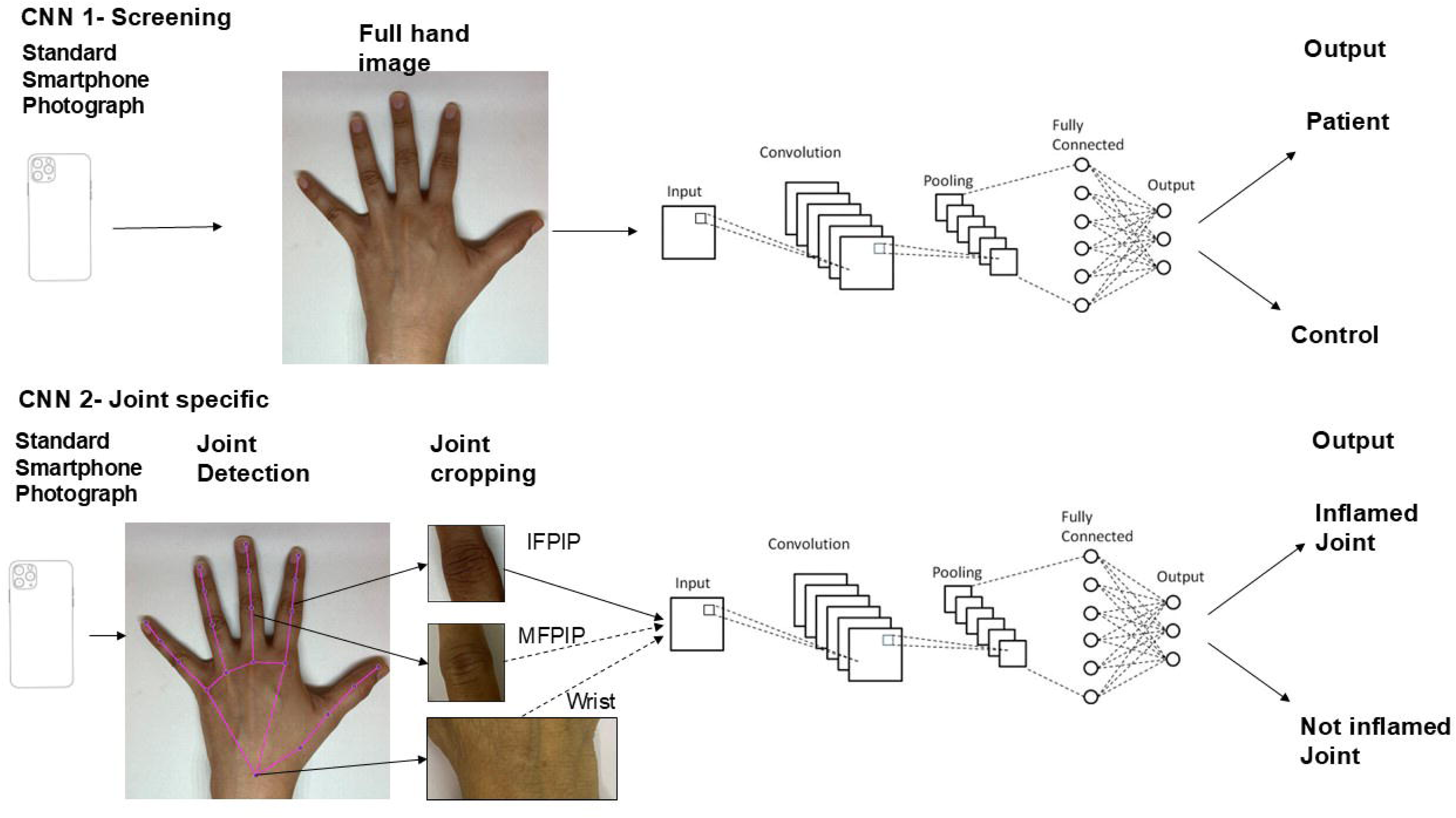
Schema of workflow to deploy image processing and convolutional neural networks (CNNs) on smartphone photographs for inflammatory arthritis detection; 1A: using a screening CNN on uncropped photos and 1B: Using joint specific CNNs on cropped photos.

CNN 2: Inception-ResNet-v2 CNNs were trained separately on cropped photos of three joints: Middle finger Proximal Interphalangeal (MFPIP), Index finger PIP (IFPIP) and wrist. The control group in joint specific evaluations include both healthy individuals as well as patients in whom that joint was not swollen. (Fig 1B)

Images were trained with augmentations including a random rotation between -20 and 20 degrees, a random X and Y translations of -20 to 20, a random scale between 0.8 and 1.2 and allowing random X-reflection. Gradient descent was carried out using the solver “adam” with batch sizes between 48 and 64. Neural networks were trained using the Deep Learning Toolbox in MATLAB (MATLAB version R2024a, 2024. The MathWorks Inc, Natick, Massachusetts)

### Statistics

Patient data, especially synovitis distribution, are presented as frequencies, mean (standard deviation) for normally distributed continuous variables, and median (IQR) for those that were not normally distributed. Normality was ascertained using the Shapiro– Wilk test. Agreement between two examining rheumatologists in the test set was calculated as a Cohen Kappa. We report accuracy, sensitivity, and specificity of CNN 1 in correctly classifying a whole hand image as belonging to a patient or healthy control within the validation set. Similar statistics were calculated for the detection of synovitis in the three individual joints within the validation set by the joint specific CNNs. The rheumatologist’s opinion was considered the gold standard for this analysis. Sensitivity was calculated as (a/a + b), viz true positive fraction amongst all patients with disease. Specificity was calculated as (c/c + d), viz true negative fraction in all patients without disease. Accuracy was the overall probability of a correct classification (Sensitivity × Prevalence + Specificity × (1 − Prevalence)). We calculated positive and negative predictive values, using the prevalence rates of inflammation for each joint from the entire dataset. Representative values are reported since each training run is slightly different by stochastic gradient descent.

### Ethics

This study received ethics permission from the KEM Hospital Research Centre Ethics Committee (KEMHRCEC/2018) and a waiver from the IISER Ethics Committee for Human Research (IECHR/Admin/2021/006). A data sharing agreement has been signed between the institutions for this study. All patients signed an informed consent document with special permission for the storage of photographs in the photo repository for 30 years. This study has been registered with the Clinical Trials Registry of India (CTRI/2020/08/027129).

## Results

Patients: 200 consecutive (143 female, mean age 49.6 (12.7) years) patients with inflammatory arthritis were included. The mean weight and height were 67.0 (13.5) kg and 160.0 (11.3) cm respectively. Normal distribution was observed in all the above variables. The median Erythrocyte sedimentation rate (ESR) was 36.0 (IQR 31.8) mm/h and median C-reactive protein was 9.0 (IQR 13.6) mg/dL. The patients were classified by the treating rheumatologist as follows: 134 had RA, 16 had peripheral spondyloarthritis, six had PsA, 15 had connective tissue diseases (three with SLE, four with SS, two with overlap connective tissue disease and 8 with undifferentiated or mixed connective tissue disease); there was one patient each of DM, FM, PAN, JIA and PMR. Seven patients had chronic post-viral arthritis. Thirty-one patients had synovitis in only one hand, while 124 had bilateral disease. Of the 400 patient hands, 72 had one swollen joint, 58 had two swollen joints, 58 had three swollen joints, and 91 had polyarthritis (four or more). The most common joint to be swollen was the wrist (43.2% hands) followed by the MFPIP (33.5%) and IFPIP (32.0%). DIP joint swellings were relatively rare. As patients with deformities were excluded, all patients were able to lay their hands flat inside the photo box. Both rheumatologists had good agreement in the detection of synovitis in 404 joints, with a concordance of 0.89 and Cohen’s kappa of 0.64. Disagreements were seen together and then agreed upon to make a final dataset.

Control: We enrolled 200 controls with no hand pain (129 female, mean age 37.8 (13.1) years, mean weight 64.4 (12.0) kg, mean height 162.2 (12.3) cm). None of these had any hand injuries or deformities.

### Full hand CNN performance

The whole-hand CNN achieved excellent accuracy (98%), sensitivity (98%) and specificity (98%) in predicting a patient as compared to control (representative image, fig 2 A). Considering this dataset with 48% prevalence (200/416), this had a PPV and NPV of 98.7% and 98.8% respectively.

**Figure 2:**
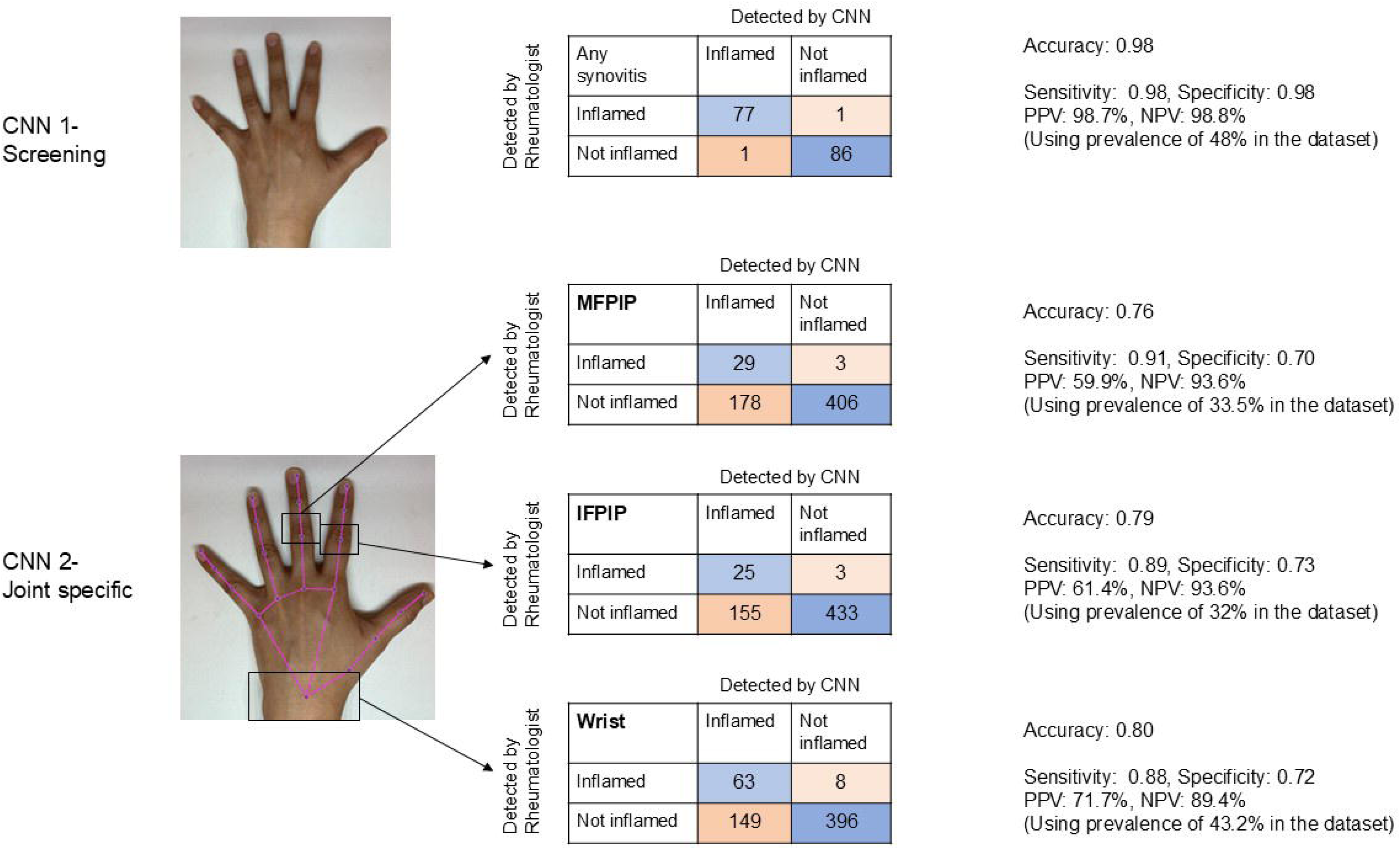
Representative images for performance of Convolutional Neural Network (CNN) for the uncropped hand photo (2A) and for three joints (2B) in contingency tables. Accuracy denotes the overall probability that a patient is correctly classified. MFPIP, middle finger proximal interphalangeal joint; IFPIP, index finger proximal interphalangeal joint; PPV, positive predictive value; NPV, negative predictive value.

Cropped joint CNN: Joint-specific CNN accuracy, sensitivity and specificity were highest for the wrist (80%, 88%, 72%) followed by the IFPIP (79%, 89%, 73%) and MFPIP (76%, 91%, 70%), fig 2B.

## Discussion

We expand on previous proof of concept results to demonstrate the ability of computer vision technologies to detect inflammatory arthritis on standardized smartphone photographs of the hands. A screening CNN could classify individuals as patients or controls with excellent accuracy. Joint specific CNNs continued to demonstrate reasonable accuracy in detecting inflammatory arthritis in three hand joints. Our results lend further credence to the possible utility of computer vision in aiding remote consults for patients with suspected or diagnosed inflammatory arthritis.

Computer vision algorithms pick up morphological signals within an image. These signals may or may not have been noticed by human eyes. Computer vision therefore promises to revolutionize fields of medicine that depend on visual data such as pathology and radiology. Supervised training on large amounts of data has allowed domain specific models such as the UNI and CONCH in computational pathology. (16) Even within rheumatology, most applications for computer vision have dealt with radiological and pathology data. These efforts have been able to automate tasks such as counting joint erosions in hand MRI in inflammatory arthritis (17) and grading the severity of osteoarthritis in knee joint X rays. (18) These advances help in research studies and clinical monitoring. In comparison, use cases of AI on point-of-care imaging, such as photographs, are relatively rare. Thermal camera images are promising: a custom CNN could differentiate RA from control based on thermal images of the hands with excellent accuracy. (9)

All these imaging technologies have standard platforms and guidelines; photographs, while easy to capture with a cellphone, suffer from a lack of standardization. Image quality and changes in background, light and equipment have been known to impact CNN performance leading to incorrect responses, as it may inadvertently learn to associate specific signals outside the object to responses. (19) We have tried to circumvent this problem by using the same smartphone for all, as well as controlling background, light and distance using a specially designed physical box. This increases confidence in the analysis but at the same time, limits application in the community. As more AI tools are used, the quality of data is likely to be important in their utility. (20) Our approach of image standardization is easy and inexpensive, and has the potential to democratize diagnosis and follow up from specialists in rheumatology to community practitioners, while also aiding a more robust follow up record in the form of stored photographic histories. Various groups have attempted to study telemedicine in these chronic disorders to help save travel resources and efforts. The TELERA study (ongoing) will systematically evaluate the efficacy of using app based patient reported outcomes. Here again, joint count is supplied by the patient. (21) Computer vision has the potential to bring specialist-level objectivity to these evaluations.

In this analysis, we tried a new strategy of training and testing a CNN on uncropped photographs of the hand. We found that this CNN achieved a remarkable sensitivity as well as specificity in distinguishing patients from controls. Due to the ‘black box’ nature of the technology, we can only conjecture that those with inflammatory arthritis had changes in colour and contour, which was detected by the algorithm. We acknowledge that age is a possible confounder: healthy controls were, on average, younger by 10 years than the patients. This skew possibly resulted from our exclusion of other chronic metabolic diseases as well as those with osteoarthritis. However, this may have led to the CNN using skin wrinkles or other effects of aging in distinguishing patients from controls. An age and gender matched control group would, in future, be able to rule out this possibility.

MFPIP and IFPIP images included not only images from control subjects (negative for synovitis) but also patients who had both synovitis of that joint and those that did not. The trained CNN results show that synovitis was detected well among the patients but that same criterion presumably conservatively assigned synovitis to control subjects as well. We believe that providing control subject information – say using another CNN such as the whole-hand network – explicitly to the networks before they are asked to make a joint-specific evaluation can be an effective leverage in reducing (control) false positives. Exploring such patterns is left for future work.

The whole-hand CNN was highly sensitive in picking up inflammatory arthritis. This evaluation also showed it was specific, but this will need to be taken with caution since ‘false positives’, viz non articular inflammatory disorders or non-inflammatory arthritis such as osteoarthritis were not compared against. Highly sensitive tests are useful as screening, before being subjected to more specific tests for confirmation (22) We envisage that a typical patient with hand pain in the community would be screened by an uncropped image CNN first. Depending on the joint involved, a joint specific CNN could be used to confirm the presence of inflammatory arthritis. Such a two step testing process is commonly used in clinical medicine, for example: screening for autoantibodies with a sensitive immunofluorescence test, followed by applying a specific test for a suspected autoantibody using enzyme linked immunosorbent assay (ELISA) or western blot. (23) Our results can allow case simulations using Fagan nomograms in assessing the diagnostic value of these algorithms in making a diagnosis. (24)

This paper has several key strengths: Firstly, the inclusion of a patient group with a diverse set of diseases, along with internal controls and healthy controls, allows for a comprehensive assessment of the model’s ability to distinguish arthritis. The patient group mimics what is seen in the clinic. Additionally, the strategy of using a screening algorithm followed by joint-specific algorithms could effectively mimic a clinical decision-making process, and also reduce the problem of false positives that the joint specific algorithm faced. Moreover, the use of standardized photographs with homogenous backgrounds and lighting conditions improves confidence that the model is not influenced by extraneous visual factors.

Our major limitations include the relatively small and ethnically homogenous dataset, which may constrain the model’s generalizability to a broader population. Additionally, the focus on only three joints limits the scope of the model, although we believe that it will behave similarly on other PIPs, subject to testing with larger datasets. The control group did not include individuals with hand pain, degenerative arthritis, but future work in this ongoing collection will assess the accuracy in distinguishing synovitis from these differentials. The reliance on a rheumatologist’s opinion as the gold standard, as compared to verifiable measures like MRI imaging, may introduce subjectivity and variability in the diagnosis. Indeed, as the less-than-perfect agreement in two rheumatologists demonstrated, it is indeed subjective. We tried to bridge this by using, in a subset, a consensus with multiple human raters; secondly-the aim of this evaluation is to bring an average specialist level examination into the community rather than detection of very early synovitis on MRI. Lastly, the absence of a formal Bayesian analysis to link the two CNNs leaves a gap in understanding how the outputs of the screening and joint-specific algorithms interact and contribute to the final diagnostic decision.

Our future plans for advancing the use of such CNNs involve expanding to a larger and more ethnically diverse dataset, allowing for greater generalizability and detection accuracy in real-world scenarios where the prevalence of arthritis is lower. We aim to refine our models by including additional joints and developing strategies for formally linking the two CNNs to enhance arthritis detection.Incorporating long-term follow-up will help validate the model’s sensitivity to change. Finally, adopting a multi-modal approach, integrating other parameters such as patient history and investigations, will help create a more comprehensive diagnostic tool that leverages computer vision.

## Data Availability

All data produced in the present study are available upon reasonable request to the authors

## Funding

The author(s) declare that no financial support was received for the research, authorship, and/or publication of this article.

## Conflict of interest

SP and PG are coinventors on a provisional patent application at the Indian Patent Office (awaiting examination for final) that includes some of the material used in this manuscript. SP is co-founder at Med2Measure Pvt. Ltd, that deals with remote detection of arthritis.

The remaining author declares that the research was conducted in the absence of any commercial or financial relationships that could be construed as a potential conflict of interest.

## Data availability statement

Anonymised photograpghs and non identifiable clinical information are available with the authors. This will be made available upon request.

## Author contributions

SP: Conceptualization, Data curation, Formal analysis, Project administration, Resources, Writing – original draft, Writing – review & editing. RC: Data curation, writing-original draft. VS: Data curation, writing-original draft, Writing-review and editing. RS: Data curation. AZ: Data curation, writing-review and editing. PH: Data curation, project administration. SC: Data curation, Formal analysis, Writing – original draft. PG: Conceptualization, Formal analysis, Methodology, Software, Writing – original draft, Writing – review & editing.

## Acknowledgments

SC is currently supported by the University Grants Commission, Government of India. We thank staff nurses in the KEM Hospital for help in data collection.

---

**Table 1:** Distribution of synovitis in the hands in the cohort, on rheumatologist’s opinion.

| **Swollen Joints** | **Left hand**<br>**(n=200)** | **Right hand**<br>**(n=200)** | **Total**<br>**(n=400)** |
| --- | --- | --- | --- |
| **Wrist** | 84 | 89 | 173 (43.2%) |
| **MCP1** | 11 | 24 | 35 (8.75%) |
| **MCP2** | 26 | 38 | 64 (16%) |
| **MCP3** | 27 | 37 | 64 (16%) |
| **MCP4** | 6 | 8 | 14 (3.5%) |
| **MCP5** | 7 | 6 | 13 (3.2%) |
| **PIP1** | 26 | 31 | 57 (14.2%) |
| **PIP2** | 61 | 67 | 128 (32%) |
| **PIP3** | 61 | 73 | 134 (33.5%) |
| **PIP4** | 28 | 30 | 58 (14.5%) |
| **PIP5** | 23 | 31 | 54 (13.5%) |
| **DIP2** | 3 | 3 | 6 (1.5%) |
| **DIP3** | 3 | 3 | 6 (1.5%) |
| **DIP4** | 3 | 2 | 5 (1.2%) |
| **DIP5** | 3 | 3 | 6 (1.5%) |
| **No swollen joints** | 59 | 59 | 118 (29.5%) |

MCP= metacarpophalangeal, PIP: Proximal interphalangeal, DIP: Distal interphalangeal

## Notes

### Funding Statement

This study did not receive any funding

### Author Declarations

Ethics committee of KEM Hospital Research Centre gave ethical approval for this work.

